# Forensic genomics of a novel *Klebsiella quasipneumoniae* type from an NICU in China reveals patterns of genetic diversity, evolution and epidemiology

**DOI:** 10.1101/2020.03.07.20032706

**Authors:** Laura Perlaza-Jiménez, Qing Wu, Von Vergel L. Torres, Xiaoxiao Zhang, Jiahui Li, Andrea Rocker, Trevor Lithgow, Tieli Zhou, Dhanasekaran Vijaykrishna

## Abstract

During March of 2017 a neonate patient suffered severe diarrhea and subsequently developed septicemia and died, with *Klebsiella* isolated as the causative microorganism. Coincident illness of an attending staff member and three other neonates with *Klebsiella* triggered a response, leading to a detailed microbiological and genomics investigation of isolates collected from the staff member and all 21 co-housed neonates. Multilocus sequence typing and genomic sequencing identified that the *Klebsiella* from all 21 neonates was a new MLST ST2727, and belonged to a less frequently detected subspecies *K. quasipneumoniae* subsp. *similipneumoniae* (KpIIB). Genomic characterization showed that the isolated ST2727 strains had diverged from other KpIIB strains at least >90 years ago, whereas the neonate samples were highly similar with a genomic divergence of 3.6 months and not related to the staff member, indicating that transmission did not occur from staff to patient or between patient to patient, but were acquired from a common hospital source. The genomes revealed that the isolates contained the ubiquitous *ampH* gene responsible for resistance to penicillin G, cefoxitin and cephalosporin C, and all Kp-IIB strains were competent for host cell adhesion. Our results highlight the clinical significance and genomic properties of relatively mild, but persistent MLST types such as ST2727, and urges for genomic surveillance and eradication within hospital environments.

**Data summary:** Genome sequences generated in this study are available in NCBI under BioProject ID PRJNA610124. All bioinformatic protocols used to process the genomic data are available at https://github.com/vjlab/KpIIB_ST2727.

## INTRODUCTION

Species of *Klebsiella* are widespread in the environment, found in soil and ground-water, as commensal organisms on plants and are carried widely in animal hosts (1). *Klebsiella pneumoniae*, is recognized as a leading cause of healthcare-associated urinary tract infections, surgical site infections and pneumonia, increasingly threatening neonates and the immunocompromised (2). However, in recent times, an increasing diversity of *Klebsiella* species have been identified to cause infection in humans (3, 4). Genome sequencing has revealed that bacteria historically classified as *K. pneumoniae* belonged to the *K. pneumoniae* species complex containing multiple phylogenetically distinct but closely related bacterial species. These include *K. pneumoniae* (previously classified as phylogroup KpI), *K. quasipneumoniae* (subsp. *quasipneumoniae* (KpIIA) *and* subsp. *similipneumoniae* (KPIIB)), *K. variicola* (KPIII) (5), and two strains forming divergent lineages indicating potentially novel species within this species complex (6). Whole genome sequencing (WGS) has shown that the predominant cause of clinical infection worldwide has been due to KpI (5), although there is an increasing trend, and increasing concern, in the detection of severe cases due to the newly described species (7). Different studies report *K. quasipneumoniae* isolates causing infections resistant to treatments with ceftazidime and other oxyimino-betalactam antibiotics, and presenting carbapenemase (KPC) genes (3, 4, 8-11).

Due to the emergence of a greater diversity of *Klebsiella* causing severe infections, their ability of colonizing environmental niches such as sinks and ventilators in hospitals (12), and increased detection of hybrid strains with drug resistant and hypervirulent phenotypes, especially in regions such as China where the earliest reports of hvKp were observed about three decades ago (13), an early warning system for the detection of *Klebsiella* species is promoted in hospitals in China (14). Routine surveillance is conducted using Multilocus Sequence Typing (MLST), followed by antimicrobial and virulence testing providing an avenue to investigate potential outbreaks and develop better control health measures.

Here we report a retrospective study of an incident that occurred in March 2017, in the NICU of a major hospital with >3000 beds in China. A neonate patient suffered severe diarrhea and subsequently developed septicemia and died, with *Klebsiella* isolated as the causative microorganism from a blood sample. During the same period, severe symptoms of diarrhea were found in three other neonate patients as well as an attending NICU staff member, triggering a response to quell the apparent outbreak. In addition to intensified hygiene procedures, the response included MLST and microbiological AMR screening of stool samples from patients (21 neonates) and the staff member. Preliminary microbiological characterization of the samples revealed that 20 of the isolates taken from the infants belonged to a new MLST designation ST2727 and one infant was infected with ST477. The sample from the staff member was of ST23 type, a well-defined hypervirulent clonal group frequently detected in severe cases of infection, whereas little was known of ST2727 and ST477. To forensically dissect the details of this case, we generated whole genome data for all bacterial isolates and carried out a detailed genomic characterization of virulence and antimicrobial resistance. Using a globally sampled dataset comprising 3611 genomes we elucidate their genetic diversity, evolution and epidemiology illuminating elements of nosocomial *Klebsiella* transmission and ecological colonization that impact generally on understanding hospital-acquired infections.

## METHODS

### Collection of clinical isolates and clinical data

*Klebsiella* was isolated from the blood sample taken prior to death of a neonate, and stool samples were collected from all 21 infants in the NICU and a staff member that suffered an episode of enteritis. These isolates were characterized using a Vitek II system (bioMérieux, Marcy l’Etoile, France). Relevant clinical characteristics of the patients and staff were extracted from their medical records, including demographic characteristics, underlying medical conditions, clinical manifestations, treatments and outcomes. This retrospective study was designed in accordance with the Declaration of Helsinki, 2013 (15) and been approved by the First Affiliated Hospital of Wenzhou Medical University (China).

### Molecular genotyping and sequence typing of isolates

Bacterial isolates were characterized by pulsed-field gel electrophoresis (PFGE) after digestion of genomic DNA samples with XbaI under the following conditions: temperature 14°C, voltage 6 V/cm, pulse angle 120°, and pulse duration of 5-35 seconds for 18h, using a well-characterized XbaI digest (Takara Bio, Inc. Japan) of *Salmonella enterica* serotype H9812 as molecular marker. DNA digest patterns were analysed and interpreted according to Tenover *et al*. (16). Multilocus sequence typing (MLST) was carried out by PCR amplifying seven housekeeping genes: *gapA, infB, mdh, pgi, phoE, rpoB* and *tonB* (17), and the data used to assign sequence types (STs) using the Institute Pasteur MLST database (https://bigsdb.pasteur.fr/). Through this process, the new ST2727 was established. Additionally, a standard multiplex-PCR was used to confirm that the isolates did not contain any of the five virulent extracellular polysaccharide capsules (CPS) (K antigen status; K-type) K1, K2, K20, K54 and K57.

### Bacterial genome sequencing and assembly

DNA was extracted using a Biospin Bacterial Genomic DNA extraction kit (Hangzhou Bioer Technology Co. Ltd.). Libraries were prepared using the TruePrepTM DNA Library Prep Kit V2 for Illumina (Vazyme). Briefly, the DNA sample was fragmented and tagged with adapters using a single transposase enzymatic reaction, followed by amplification using an optimized, limited-cycle PCR protocol and indexing. Individual libraries were assessed on the QIAxcel Advanced Automatic nucleic acid analyzer, and quantitated through qPCR using KAPA SYBR FAST qPCR Kit. Illumina Next Generation Sequencing technology (Hiseq PE150) was used to generate a paired-end library of short read sequences (150nt) for each isolate. Sequence quality controls were performed using FastQC version 10-01-18 (Andrews, n.d.) and trimmed and refined using Fastx toolkit version 0.0.13 (Lab, n.d.). *De novo* assembly was performed using SPAdes version 3.13.1 (18). Scaffold generation and ordering were performed using MeDuSa version that was available on July 2018 and Mauve version 2.4.0, respectively (19, 20). Closely related genomes (NTUH-K2044 (Kp9), CP027602 (Kp10), HKUOPLC (Kp1-8, Kp11-22) from NCBI were identified using genome identity. Upon assembly and alignment of all ST2727 genomes, single nucleotide variations (SNV) were inferred using Parsnp tool from Harvest Suite version 1.1.2 using a reference Kp1 isolate genome (21).

### Genomic analysis

Genes contributing to drug-resistance and virulence were detected using Kaptive version available on July 2018 and Kleborate version 0.3.0 (22, 23). VRprofile version 2.0 was used to detect genes clusters characteristic of protein secretion systems of type 3 (T3SS), type 4 (T4SS) or type 6 (T6SS) (24). The presence of genes encoding a type 2 secretion system (T2SS) was detected by BLAST version 2.7.1, using as queries the known T2SS genes (25). Phylogenetic analysis was conducted using 3611 *Klebsiella* genomes downloaded from the NCBI database (downloaded on 20/06/2018) to find the closest related genomes to the novel strains (Data S1). Core genome alignment of *Klebsiella* genomes (including ST2727 isolates) was generated using Roary version 3.11.2. Excluding poorly assembled genomes the final dataset constituted 3611 genomes comprising 3251 genes that were in at least 99% of the sequences (26). Phylogenetic relationships were estimated using the maximum likelihood (ML) method in RAxML version 8.2.12 (27) using the General Time Reversible (GTR) nucleotide substitution model with a gamma (Γ) distribution of among-site rate. Branch support was estimated using a ML bootstrap analysis with replicates ranging from 10 to >1000 replicates for the different datasets analysed.

Finally, to detect signatures of natural selection in the ST2727 outbreak samples, we used McDonald-Kreitman Test (MKT) implemented in the Rpackage PopGenome (28). MKT was calculated for the whole genome, and gene-wise. The MKT is the ratio (neutrality index, NI=*(Pn/Ps)/(Dn/Ds)*) of the relation of nonsynonymous/synonymous mutations between strains (*Pn/Ps*) and the relation of nonsynonymous/synonymous mutations within strains (*Dn/Ds*). When this ratio is 1 it means that the same number of mutations are happening between and within strains, which suggests that there is not selective pressure or neutral selection occurring within strains. When the ratio is more than 1 (NI>1) it means that *Pn/Ps>Dn/Ds*, which is characteristic of negative selective pressure occurring within strains. The opposite, a ration less than 1 (NI<1), when *Pn/Ps<Dn/Ds*, is characteristic of positive selection occurring within strains (29).

To confirm that the genome datasets contained sufficient genetic change between sampling times, necessary for the reconstruction of the time-scale of evolution, we used a root-to-tip regression of sampling years against genetic diversity in TempEst v1.5 (30), optimizing the best fit for the root to maximize the determination coefficient, *R*^2^. The slope of the regression was positive, showing that the genomic data reflects temporal signal, so a molecular clock was estimated using a least-squares dating (LSD version 0.3) method (31) with 1000 samples for the confidence interval. The reliability of the analysis was confirmed using a permutation test in which the sampling years were randomized for 100 replication, with a Zscore test (32). The nucleotide substitution rate per year and the branch lengths were used to determine the time of divergence of each node.

## RESULTS

### Clinical scenario and diagnosis

This study was initiated when outbreak status was called in a NICU due to a fatal case of a neonate patient presenting with symptoms of respiratory and gastrointestinal infections. Despite treatment with meropenem, vancomycin, cefoperazone and sulbactam, the neonate patient died from neonatal septicemia, septic shock, gastrointestinal hemorrhage and multiple organ dysfunction syndrome. Days after the fatal case, 20 more neonates were sampled along with an NICU staff member who presented symptoms of infection (Fig 1C). According to a Vitek II system (see Methods), blood and stool samples collected from the 21 neonates co-housed in the single-room NICU and the staff member were positive for *K. pneumoniae*. The neonates presented with a wide range of symptoms: three with respiratory and digestive symptoms, thirteen with respiratory disease such as dyspnea, severe asphyxia, tachypnea and/or cyanosis severe, one with digestive symptoms alone, while three presented with normal health (Table 1). The staff member presented with digestive symptoms including vomiting and severe diarrhea. MLST type classification using the *gapA, infB, mdh, pgi, phoE, rpoB* showed that 20/21 neonate isolates were novel with a new MLST designation ST2727, whereas one neonate was of ST477 and the staff member was infected with a well-characterised hypervirulent type ST23, suggesting that the fatality was caused by the ST2727, and demonstrating that the staff member was not the cause of the event. Here, we sought to understand the carriage and genomic characteristics of ST2727 in the neonates and test the hypothesis of transmission in this closed scenario.

**Table 1.** Patient and *Klebsiella pneumonia* strains.

| Sample id | MLST<sup>1</sup> | Patient | Birth | Symptoms<sup>2</sup> | Antibiotics | Invasive procedure |
| --- | --- | --- | --- | --- | --- | --- |
| KP1 | ST2727 | Neonate | vaginal | Digestive, Respiratory | yes | Yes |
| KP2 | ST2727 | Neonate | cesarean | Digestive, Respiratory | yes | Yes |
| KP3 | ST2727 | Neonate | cesarean | Digestive, Respiratory | yes | Yes |
| KP4 | ST2727 | Neonate | vaginal | Respiratory | no | Yes |
| KP5 | ST2727 | Neonate | vaginal | Respiratory | no | Yes |
| KP6 | ST2727 | Neonate | cesarean | Respiratory | yes | Yes |
| KP7 | ST2727 | Neonate | cesarean | Respiratory | yes | Yes |
| KP8 | ST2727 | Neonate | cesarean | Respiratory | no | No |
| KP9 | ST23 | Staff | N/A | Digestive | - | - |
| KP10 | ST477 | Neonate | vaginal | Digestive | yes | No |
| KP11 | ST2727 | Neonate | cesarean | Respiratory | yes | Yes |
| KP12 | ST2727 | Neonate | vaginal | Respiratory | yes | No |
| KP13 | ST2727 | Neonate | cesarean | Asymptomatic | no | no |
| KP14 | ST2727 | Neonate | cesarean | Respiratory | yes | Yes |
| KP15 | ST2727 | Neonate | cesarean | Respiratory | no | No |
| KP16 | ST2727 | Neonate | vaginal | Respiratory | no | Yes |
| KP17 | ST2727 | Neonate | cesarean | Respiratory | yes | Yes |
| KP18 | ST2727 | Neonate | vaginal | Noninfection | no | Yes |
| KP19 | ST2727 | Neonate | vaginal | Asymptomatic | no | No |
| KP20 | ST2727 | Neonate | vaginal | Respiratory | yes | Yes |
| KP21 | ST2727 | Neonate | cesarean | Respiratory | yes | Yes |
| KP22 | ST2727 | Neonate | cesarean | Asymptomatic | no | No |

<sup>1</sup>Multilocus Sequence Typing. <sup>2</sup> Symptoms are classified depending of what type of infection they are related to, “noninfections” suggests the patient has symptoms related to other diseases that are non-infectious disease for example symptoms related with cardio-respiratory chronic diseases, “Asymptomatic” refers to no symptoms at all. “Respiratory” and “Digestive” refers to symptoms related to respiratory infections and digestive infections, respectively.

**Figure 1.**
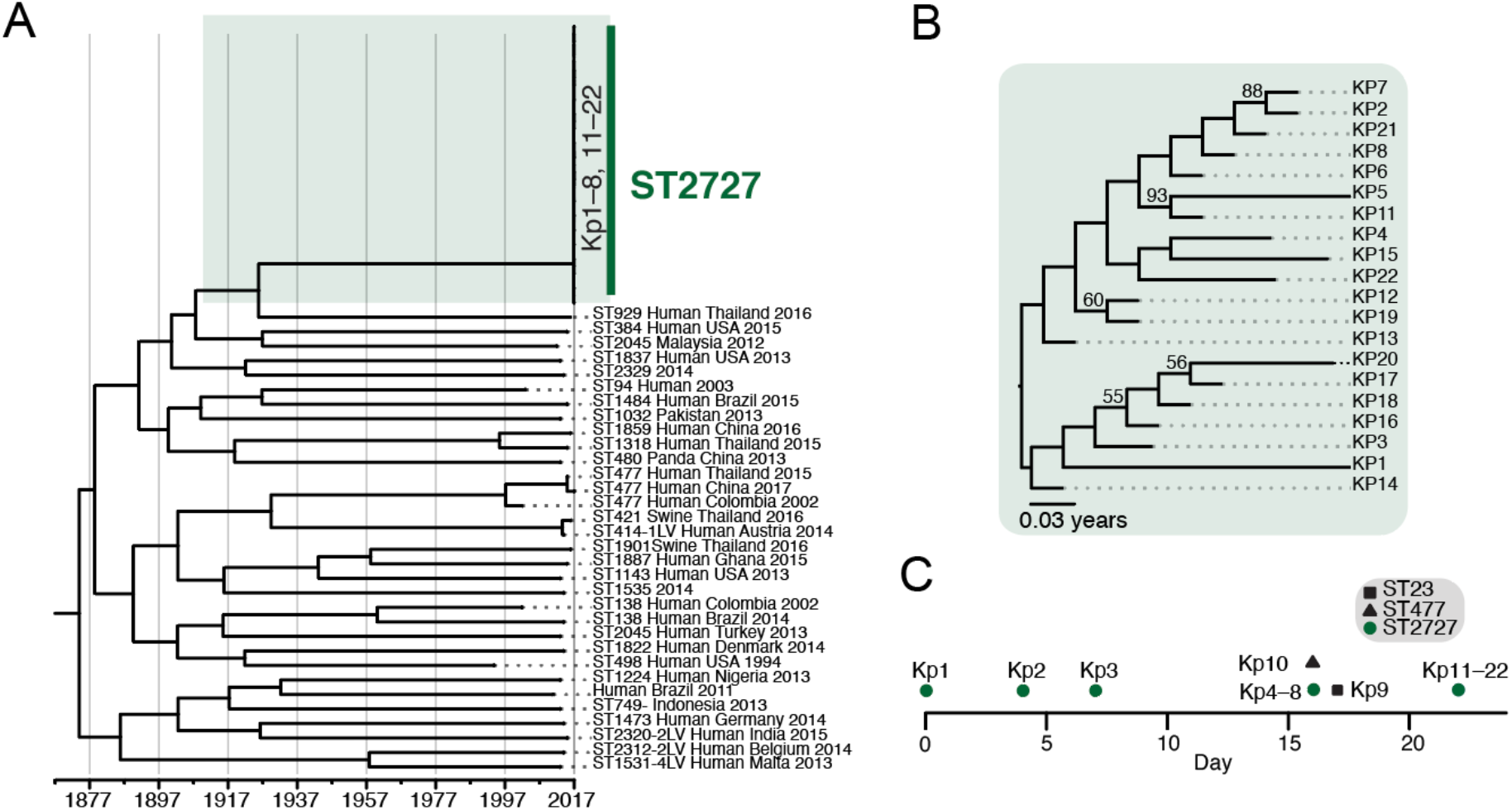
Genomic epidemiology and timeline of *K. quasipneumoniae* subsp. *similipneumoniae* (KpIIB) sample collection in NICU. (A) Evolutionary relationships and time-scale of evolution of 54 *K. quasipneumoniae* subsp. *similipneumoniae* (KpIIB) strains, and (B) ST2727 isolates detected in NICU during February and March 2017, generated using the least-square dating method. (C) Timeline of sample collection in NICU, where the calendar dates have been replaced by a timeline where the first sample was collected during day 0.

### Genomics-based evaluation of the NICU strains

To determine the genomic characteristics of the bacteria recovered in this study, the complete genomes of all 20 ST2727 isolates were sequenced using a paired-end short read sequencing methodology. The lengths of the assembled genomes ranged from 5,192,865 bp to 5,202,717 bp that encoded between 4706 and 5191 annotated genes (Table S2). The GC content of all genomes was 57%. Genomic analysis showed the isolates exhibited high genomic similarity (0.018% sequence divergence), with 1,066 single nucleotide variations (SNV) along their genomes corresponding to an average of one SNV every 2,392 bases. The variation observed among the ST2727 samples were at the lower range to the median divergence of 0–0.08% observed among the same MLST types (5).

Phylogenies generated from a core genome of 3611 globally sampled *Klebsiella* strains (Fig S1, <u>Data S1</u>), including isolates from the present study made evident that the ST2727 strains belong to *K. quasipneumoniae* subsp. *similipneumoniae* (KpIIB) (33) (Fig. 1A, Fig S2). The high similarity of the ST2727 strains suggests a clonal population. A root-to-tip regression of sampling dates and genetic distance of 54 KpIIB genomes sampled during 1994–2017, including the ST2727 genomes, showed a positive correlation (correlation coefficient: 0.64) between time and mutation, indicating that the dataset contained sufficient signal for the estimation of a molecular clock. We used the least-square dating method, to estimate an evolutionary rate for the KpIIB clade of 4.596 × 10^−05^ nucleotide substitutions per site per year (nt subs/s/y) (95% CI of 4.180 to 4.980 × 10^−05^). The ST2727 group exhibited a genomic divergence of >90 years (mean time of most recent common ancestor (TMRCA) 1926; 95% confidence interval (95% CI), 1917 to 1933) to the most closely related isolate collected in 2016 from a patient from Thailand (KPPSTH03; MLST type ST929-1LV) (Fig 1A).

The TMRCA of all KpIIB lineages was 1874 (95%CI, 1861 – 1886), whereas, the mean TMRCA of ST2727 collected within the hospital was approximately 3 months prior to the first case Kp1 (mean TMRCA 2016.94, 95%CI 2016.88 – 2016.97) (Fig 1B). This showed a divergence time much greater than the duration of sampling across 22 days in February–March 2017 (Fig 1C). These results suggest that what was designated as an outbreak was instead the colonization of infants by a community of bacteria that were transmitted to the neonates during this period. Due to the variation present between ST2727 isolates, it is unlikely that the colonization occurred through direct transmission between one neonate to another. We suggest instead that the isolates were introduced to the hospital or the ward in the months prior to the outbreak and sustained within the hospital during this period. The precise mechanism of colonization of the neonates is not clear without comprehensive sampling of the built environment and the equipment used in treatment and care of the neonates.

The core genome of ST2727 clade exhibited positive selection in comparison with strains in the KpIIB clade (MKT, neutrality index: 0.35, NI < 1), indicating that ST2727 isolates are under selection pressure. Gene-wise estimates of selection pressure in the ST2727 clade in comparison with KpIIB strains showed that 21 genes were under negative selection (NI >1) (Table S4), 10 were under positive selection (Table S5), whereas, mutations in the remaining genes were either strongly deleterious or were under neutral selection pressure (29). Many of these genes are fundamental to bacterial cell biology and have been functionally characterized in various *K. pneumoniae* strains, and these functions can be expected to be conserved in *K. quasipneumoniae* subsp. *similipneumoniae* (KpIIB). Genes under purifying or negative selection included genes coding for transcriptional factors, metabolic enzymes, a structural protein, and proteins coding for active transport of solutes (Table S4). Transporter proteins found to be under purifying selection conform to a set of protein families: the ATP-Binding Cassette (ABC) transporters, Major Facilitator Superfamily (MFS) transporters, and antiporters and symporters. ABC transporter under negative selection were those designated as responsible for the transport of thiamine, nickel and carbohydrates, all of which are required for bacterial survival in nutrient rich environments (34).MFS transporters uptake nutrients from rich environments using chemiosmotic ion gradients (34), while antiporters and symporters maintain ion homeostasis for these and other functions (34). Taken together, this suggests that the ST2727 population has adapted for growth in a nutrient poor environment. Conversely, while the genes under positive selection also include some for active transportation of solutes (Table S5), these transporters are responsible for export of toxins and antibiotics as efflux systems. For example, genes coding for efflux RND transporter permease subunits (drug efflux) and HlyD toxin secretion (34). This reliance on drug-efflux is consistent too with the observation that the structural protein peptidoglycan glycosyltransferase/peptidoglycan DD-transpeptidase (PBP1A) encoded by the gene *mrcA*, was under negative selection: PBP1A catalyses the transglycosylation and transpeptidation of murein that is a major component of the cell wall, but it is also the target of beta-lactam antibiotics (34).

### O- and K-antigen serogroups of NICU isolates

WGS analysis using Kaptive from Kleborate (22, 35) identified that ST2727 strains have a KL55 capsular type (*wzi*56). O-typing of the surface lipopolysaccharide revealed all ST2727 were O3/O3a. This serotype has a strong adjuvant effect compared with other lipopolysaccharide types (36), but is a rarely reported serotype with no known clinical impact (37). Genome sequence data revealed that all isolates were *wcaG*– (Fig 2A).

**Figure 2.**
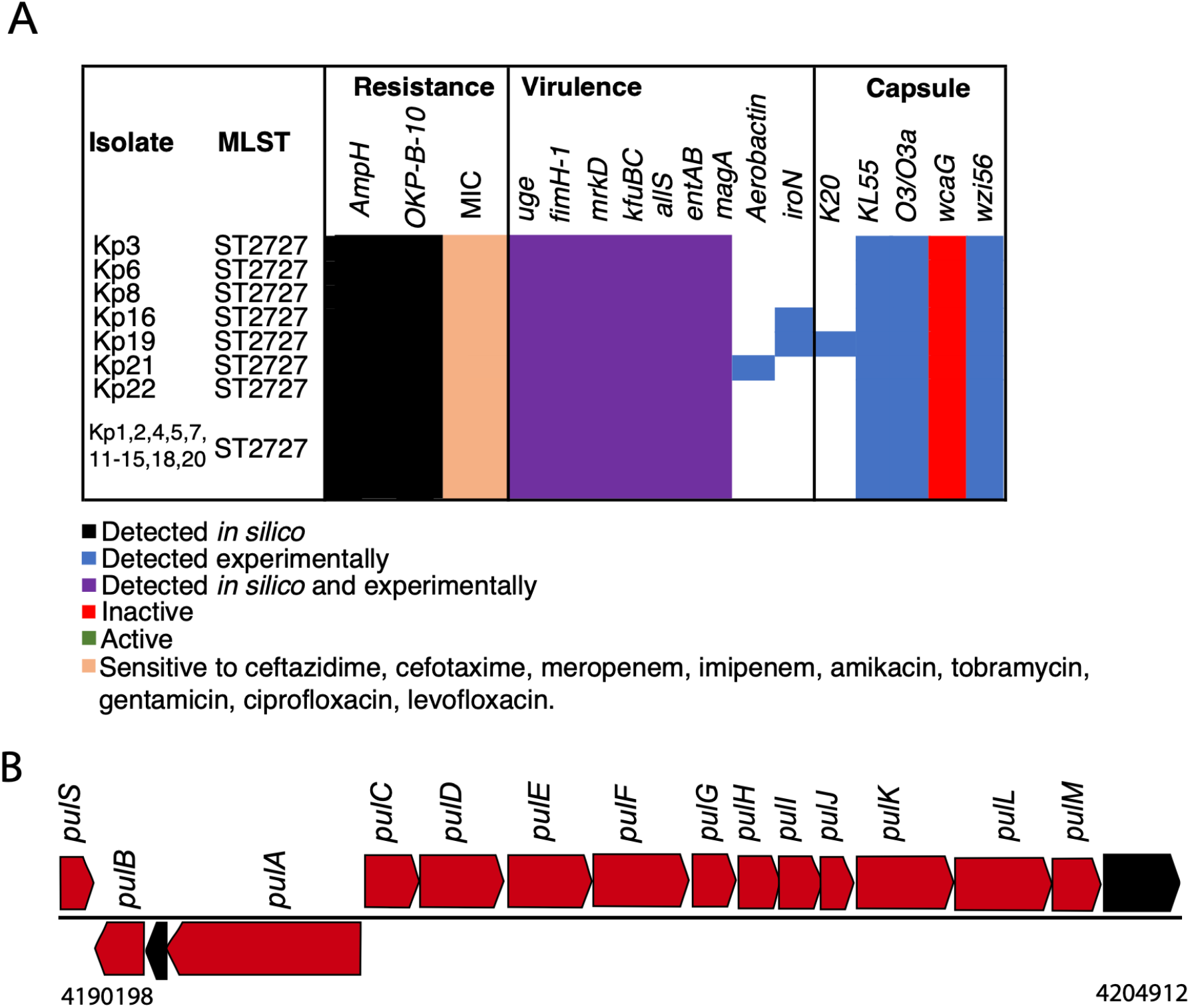
Major resistance, virulence and capsular genes detected in ST2727. (A) Summary of genes contributing to drug-resistance, iron-acquisition and other virulence-enhancing features. (B) Genomic architecture of the T2SS cassette in ST2727.

### Genes associated with virulence and antimicrobial resistance

All ST2727 isolates shared three genes considered diagnostic as virulence factors (Fig 2A, Table S3): *uge* (uridine diphosphate galacturonate 4-epimerase), *fimH* (type 1 fimbrial adhesion) and *mrkD* (type 3 fimbrial adhesion). *Uge* is responsible for the conversion of UDP-GlcA to UDP-GalA and is key for the production of polysaccharides containing GalA residues, a feature which has been shown to promote colonization of human tissues (38). Fimbriae are an important feature of *Klebsiella*, and the gene *fimH* encodes a well-conserved adhesin subunit that is found in around 90% of *Klebsiella* strains (39). This gene has high mobility within clones through horizontal transfer (39). The gene encoding *mrkD* is associated with “type III” fimbriae, important for adhesion to promote establishment of bacterial biofilms in harsh environments (40).

In terms of antimicrobial resistance, two genes encoding beta-lactamases were detected (Fig 2A, Table S3): the ubiquitous *ampH*, responsible for resistance to penicillin G, cefoxitin and cephalosporin C, and the Kp-IIB restricted *bla*OKP-B-10 (41).

### Secretion Systems

Protein secretion systems are key components of virulence in many bacterial pathogens (42), and in *Klebsiella* the type 2 secretion system (T2SS) is the best characterized of these virulence determinants (43). *In silico* inspection of the genomes showed that all ST2727 isolates collected have a similar T2SS architecture, typically composed of *pulS* and *pulA-pulM* (44) (Fig 2B). HlyD, is a diagnostic component of the type 1 secretion system (T1SS), and was detected in the ST2727 strains (Table S5). Other common secretion systems (T3SS, T4SS, T5SS and T6SS) were not found in any of ST2727 isolates.

### Evolutionary relationship of ST2727 to globally isolated *Klebsiella* strains

*Klebsiella quasipneumoniae* was previously considered of low clinical risk (3-5). Mapping of virulence and resistance phenotypes observed among *K. quasipneumoniae* subsp. *similipneumoniae* (KpIIB) and globally collected *Klebsiella* genomes showed that two KpIIB MLST types, ST1901 (GCF_002248055.1) and ST2320 (GCF_001729665.1) and a KPIII (*K. variicola*) type (ST981) (GCF_001463685.1) (45) contained virulence markers, that were previously only observed among *K. pneumoniae* strains (22, 23). The ST1910 genome collected in Thailand in 2016 contained the virulence marker aerobactin (*iuc3*), whereas the ST2320 strain collected in India during 2015 can express aerobactin (*iuc1*), yerseniabactin (*ybt 9*; *ICEKp3*) and salmochelin (*iro1*), as well as producing a hypermucoidy phenotype (*rmpA_4, rmpA2_3*). The emergence of virulence markers among KPII genomes raises concerns as there has been a rise in documentation of hospital-acquired infections caused by *K. quasipneumoniae* (5, 7). Also, Figure 3 shows that ST2727 is closely related to ST498, a group that has been found to have resistance genes including *aadB, sul1, blaOKP-B-7*, and the genes *blaOXA-2* and *blaSHV-18* that encode ESBL (8) (Fig. 3).

**Figure 3.**
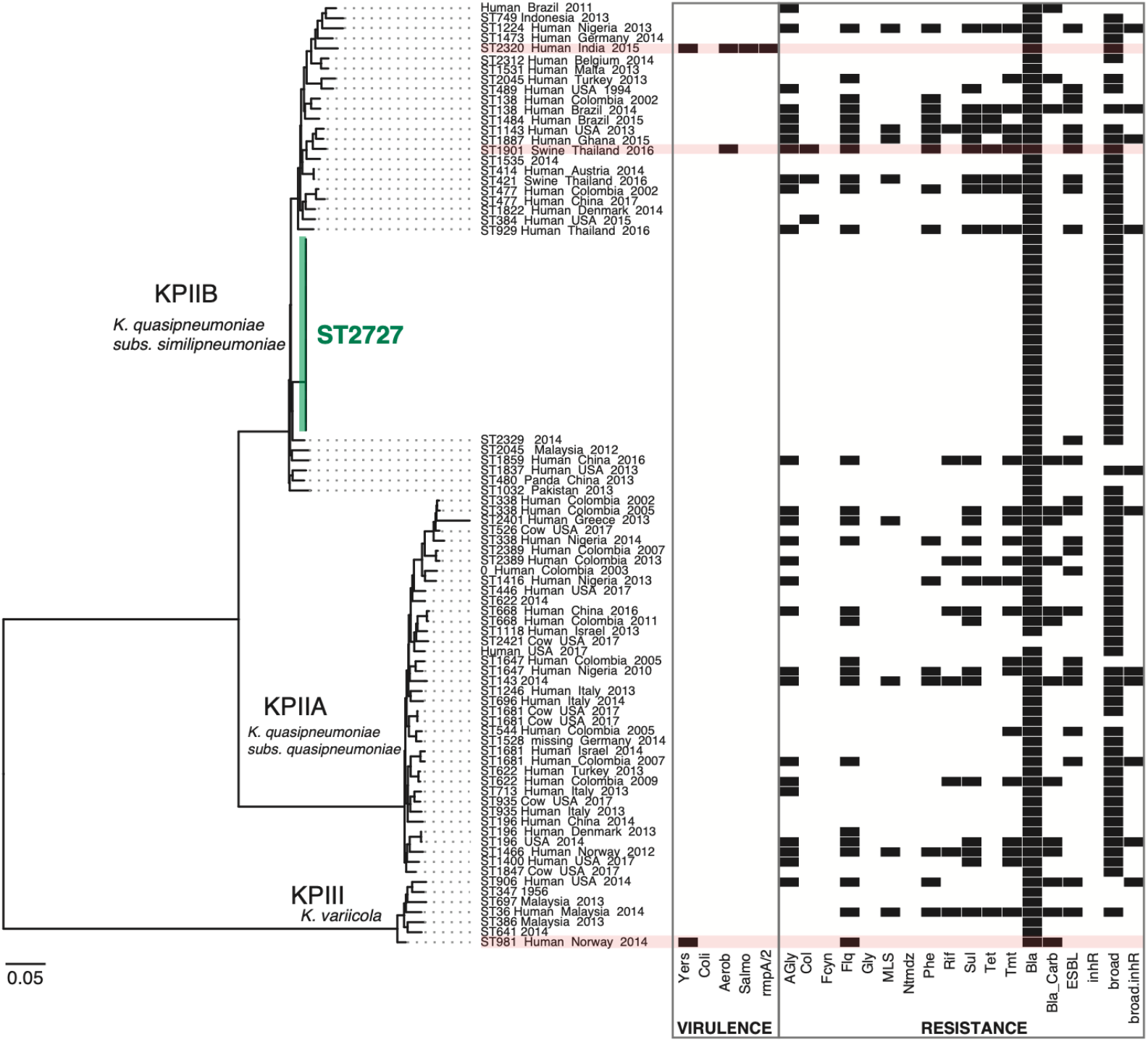
Phylogenetic distribution of virulence and resistance genes among KPII and KPIII genomes. The presence of virulent and resistance genes is shown in black. Virulent genes are grouped in categories: Yers (yersiniabactin), Coli (colibactin), Aerob (aerobactin), Salmo (Salmochelin) and *rmpA, rmpA_2* (hypermucoid). Resistant genes are grouped in categories: Gly (glycopeptides), MLS (macrolides), Phe (phenicols), Rif (rifampin), Sul (sulphonamides), Tet (tetracyclines), Tmt (trimethoprim), Bla (beta-lactamases), Bla_Carb (carbapenemase), ESBL (extended spectrum beta-lactamases), inhR (extended spectrum beta-lactamases with resistance to beta-lactamase inhibitors), broad (broad spectrum beta-lactamases), broad.inhR (broad spectrum beta-lactamases with resistance to beta-lactamase inhibitors). Virulent MLST types are highlighted in red.

## DISCUSSION

The colonization of the neonate gut is a complex process with contributions from the mother’s microbiome and environmental sources, that can not easily be disentangled (46-48). In this study, 21 neonates housed in a single room were sampled across 22 days for the carriage of *Klebsiella* in response to an outbreak-status event. It is clear from genomics data the hyper-virulent ST23 isolated from the staff member was an irrelevant coincidence to the sepsis event in the neonate patients. The only *Klebsiella* isolated from the sick neonates carried a previously unreported sequence type (ST2727), with core-genome phylogenies showing they belonged to *K. quasipneumoniae* subsp. *similipneumoniae* (33). Despite the availability of few KPIIB genomes, our analysis of 54 available genomes including the 20 new ST2727 samples, showed positive correlation between genetic distances and date of sampling (30). This indicates that our dataset was adequate to show that the NICU samples had diverged months prior to the detection of the first case, and showing that ST2727 was not transmitted between patients during the outbreak. These results were further supported by a permutation test where the sampling dates were randomized (32, 49). Furthermore, the lower sampling of KPIIB genomes, as compared to the greatly sampled *K pneumoniae* genomes, would not have an effect on the estimation of the molecular clock for the outbreak samples because bacterial phylogenies in general show a large genetic distance between MLST types – signified by the long phylogenetic branch lengths between MLST types (star-shaped species level phylogeny), indicating that adequate sampling of closely related bacteria would have an improvement in estimates rather than greater sampling of highly divergent genomes within a bacterial species. Overall, this study suggests that a population of ST2727 with some sequence variability is likely endemic within the hospital environment, and that distinct members within this population have infected the neonates in their first days of life.

It is probable that ST2727 is under antibiotic selective pressure due to their prevalence in the hospital environment for the months prior to the events analysed here. This is further confirmed with a MKT indicating positive selective pressure ongoing for genes related with efflux systems that are a key part for the transport of toxic substances as antibiotics. However, further investigation on evolutionary mechanism selecting for resistant genes and genes related with resistance mechanisms would need to be done in order to assert these findings. Previously, estimates of selection pressure on the genes encoding beta-lactamases in the KpI, KpII and KpIII clades have showed neutral selection of these genes (45). In agreement with this, we did not find either of the genes encoding beta-lactamases in ST2727 to be under natural selective pressure.

Although made highly unlikely, the data in this study cannot rule out a maternal contribution to the *Klebsiella* carriage, but we note that (i) 12/20 neonates were delivered via Caesarian section (Table 1) where maternal contribution to microflora should be inconsistent and minimal, and that (ii) ST2727 is probably not a dominant sequence type in the community as it has not been sampled previously, despite the hospital having more than 1500 *Klebsiella* isolates collected from adult patients over 15 years. We suggest that a substantial contribution to carriage comes from the endemic microflora in the NICU, with this specific built environment colonized by a population corresponding to ST2727. A suggestion for future analyses would be inclusion of more detailed samplings of the NICU environment, including the built environment itself and systematic sampling of staff members and visitors.

Previous analysis estimated that at least half of *Klebsiella* infections result from patients’ own microbiota (50). That the neonate who died likely had a bowel perforation to initiate sepsis from a strain carried in his gut supports the contention that *Klebsiella quasipneumoniae* carriage contributes to the risk of infection in hospital environments. Thus, carriage of drug-resistant or hypervirulent clones of *Klebsiella*, including *K. quasipneumoniae* in the gut would be of great concern in people with lowered immune systems – such as neonates - or those with conditions requiring surgical treatment where bowel perforation is a risk factor (46). Our study highlights the importance of WGS for elucidating the transmission and carriage of hospital-resident bacterial populations, urges for the early identification and eradication of persistent strains that increase the rate of hospital-acquired infections.

## Data Availability

All newly generated sequence data has been submitted to GenBank, and the accession numbers are included.

## ETHICAL APPROVAL

This study was approved by the Ethics Committee of the First Affiliated Hospital of Wenzhou Medical University (China).

## ACKNOWLEDGEMENTS

We thank Dr. Jiangning Song for his input in the early stages of this analysis.

